# Coverage, Utilisation and Users of a Chat-Based Digital Primary Care Clinic in Publicly Funded Healthcare: A Registry-Based Observational Study in Finland

**DOI:** 10.1101/2025.11.06.25339651

**Authors:** Alexandra Dahlberg, Taavi Kaartinen, Sakari Jukarainen, Petja Orre

## Abstract

**Background:** Chat-based digital clinics are increasingly integrated into public primary care. This study evaluates a 24/7 chat-based digital clinic serving approximately 130,000 residents in publicly funded primary care in Päijät-Häme, Finland.

**Methods:** Data from 1,580,838 primary care encounters recorded between 2019 and 2025 were used to examine care patterns following the digital clinic’s introduction.

**Results:** Primary care coverage (residents with ≥1 public primary care encounter) fell from 39.2% in 2019 to 35.7% in 2020, consistent with pandemic-related suppression, before stabilising between 44% and 48% from 2022 onwards (44.2% in 2025). Utilisation rose from 1,222 encounters per 1,000 residents in 2020 to 2,058 by 2025. Digital encounters grew from 15.2% of all primary care encounters in 2021 to 25.8% in 2025. Digital users were younger (mean age 33.2 vs. 55.1 years; *P*<.001) and had lower crude comorbidity prevalence (CCI ≥ 1: 13.1% vs. 33.6%); after age and sex standardisation this narrowed to 22.8% vs. 27.5% (adjusted OR 0.78, 95% CI 0.75-0.81). Common physician-level diagnoses included conjunctivitis, acute cystitis, and prescription renewals. Same-day escalation to a physician occurred in 18.0% of digital clinic encounters; excluding care already scheduled beforehand, 32.6% of patients had a further contact of any reason within 14 days and 42.4% within 30 days.

**Conclusions:** Primary care coverage and the digital share of encounters both rose alongside the digital clinic’s introduction. Most encounters were concluded at nurse level without a planned follow-up, and approximately 60% of seven-day pathways remained within the digital channel. Where patients contacted the service again within 30 days, a repeat digital encounter was the most common route and an in-person appointment the first return contact for fewer than one in ten patients; contacts were counted irrespective of reason.

## Introduction

Digital technologies are reshaping the delivery of primary care. Asynchronous messaging, live chat, video consultations, remote monitoring, patient portals, and structured digital care pathways can improve accessibility, timeliness, and operational efficiency [1–3]. In the Nordic context, digital primary care has been promoted both as a service innovation and as a response to rising demand, workforce shortages, and access problems in publicly funded systems. In many countries, the COVID-19 pandemic accelerated the adoption of digital care models [4,5], though interest in digital-first approaches predated the crisis [6]. In Finland, digital transformation has similarly been framed as a system-level priority, with the expectation that digital modalities can support timely access to care and alleviate pressure on outpatient services.

Chat-based digital clinics are often organised around nurse-led triage, with the aim of resolving a substantial share of care needs without physician involvement and escalating cases to physician consultations or in-person care when clinically indicated. Early implementation studies and economic evaluations suggest that such digital-first pathways can be cost-effective and may enhance system responsiveness in real-world settings [7–9].

Evidence from European settings indicates that patients using digital primary care tend to be younger and more often female than those accessing only traditional services [9–11]. Comorbidity burden among digital users has rarely been reported, and where lower prevalence has been described it has not been separated from the younger age profile of these users. Younger, predominantly female uptake raises the possibility of a digital divide, in which patients with greater health needs or barriers to access are less likely to use digital channels [12]. Yet longitudinal evidence from publicly funded systems on how integrated digital clinics affect overall coverage, utilisation, and patient pathways over time remains limited, particularly in Nordic settings.

In this study, we examine the integration of a centralised, 24/7 chat-based digital clinic into Harjun terveys, a publicly funded primary care provider serving approximately 130,000 residents in the Päijät-Häme region of Finland. Using registry-based data from 2019 to 2025, the study had four aims: (1) to describe system-level trends in primary care coverage and utilisation across a seven-year period spanning pre-implementation, early implementation, and mature implementation phases; (2) to characterise the demographic and clinical profile of digital users; (3) to describe the clinical scope of digital encounters; and (4) to examine care pathway patterns following nurse-led digital consultations.

## Methods

### Study Design, Setting and Data

Finland provides universal healthcare coverage primarily through publicly funded services, with private and occupational healthcare operating in parallel [13]. Because employed individuals typically access primary care through employer-funded occupational health services, the public primary care population skews towards children, older adults, and those outside the workforce.

Harjun terveys is a joint venture, established in January 2021, that delivers publicly funded primary healthcare to approximately 130,000 residents in the Päijät-Häme region of Finland under a multidisciplinary, team-based model with a “digi-physical” service pathway. All registered residents can access a 24/7 digital clinic providing asynchronous chat-based consultations via the Päijät-Sote application, alongside traditional telephone and in-person appointments. All patient contacts, whatever the modality, are documented in the national unified electronic health record (EHR) system. Further detail on the setting is given in Supplementary Methods 1.

This retrospective observational study used routinely collected health service data spanning 1 January 2019 to 31 December 2025, providing two pre-implementation years (2019-2020) and five years during which digital services and broader organisational reforms were introduced and scaled: system-wide digital services from January 2021, and a centralised digital clinic under a standardised model from January 2023. The period encompasses the COVID-19 pandemic, during which care-seeking and service delivery were substantially disrupted, particularly in 2020.

The extract comprised 2,902,707 pseudonymised encounter-level records, of which 2,796,395 remained after deduplication and exclusion of dental, out-of-scope, and undated records (Supplementary Methods 2). Extracted variables include patient identifier, encounter date, encounter modality, service unit, provider profession, booking status, and diagnostic codes (ICPC-2 and ICD-10). Sex, date of birth, municipality, and residence start and end dates were obtained from a corresponding population registry extract and used to define annual eligible populations and derive age at encounter. Comorbidity burden was measured using the Charlson Comorbidity Index (CCI), calculated from ICD-10 diagnostic history using an established mapping algorithm [14].

### Coverage and Utilisation

Service coverage was defined as the proportion of registered residents with at least one primary care encounter during a calendar year, and digital coverage as the proportion with at least one digitally delivered encounter. Utilisation was measured as encounters per 1,000 registered residents per year; mean annual encounters per patient was calculated as total, or digital, encounter counts divided by the number of unique patients with at least one primary care encounter that year. Population denominators were derived from national population registry data [15].

A primary care encounter was defined as an encounter with a nurse or a general practitioner delivered through the digital clinic, at a physical health centre, or as a physician consultation, whether digital or face to face; 1,580,838 encounters met this definition, including 74,845 recorded in the separate digital system. Health promotion and preventive services, telephone contacts, digital documentation entries, and encounters not classifiable to a defined modality were excluded; the rationale for each exclusion and the full record flow are given in Supplementary Methods 2. An encounter was classified as digital where the record was flagged as digitally delivered, comprising digital clinic encounters and digitally delivered physician consultations.

For 2021-2022, and for evening encounters in January-April 2023, digital clinic records were held in a separate administrative system with distinct patient identifiers that could not be linked to the primary EHR. Encounter counts, and therefore encounters per 1,000 residents and the digital share, were obtained by summing across both systems and are exact for all years. Coverage and mean encounters per patient for 2021-2023 incorporate a correction for expected cross-system patient overlap, using an empirical overlap fraction (φ = 0.654) derived from the fully linked 2024-2025 data (Supplementary Methods 3); sensitivity analyses across the full range of plausible φ values are presented in Supplementary Table S1.

### Patient Characteristics

Because pre-2023 digital clinic records lacked patient identifiers that could be linked to the EHR, all patient-level analyses were restricted to the fully linked 2023-2025 data. The unit of analysis was the person-year: each patient contributed one observation for every calendar year in which they had at least one primary care encounter meeting the definition above, and was classified as a digital user in that year if at least one of that year’s encounters was digitally delivered and as a traditional user otherwise. Patients could therefore change classification between years. Age and sex were taken from the first encounter of each year, and comorbidity burden from the population registry extract.

### Clinical Content and Care Pathways

To characterise the clinical scope of digital care, diagnoses recorded in digital clinic encounters from 2023 to 2025 were described using ICPC-2 codes for presenting symptoms and primary care-level assessments, and ICD-10 codes for physician-level digital consultations, summarised both as the most frequent individual codes and as aggregated chapters.

All contacts at the digital clinic are handled by a nurse in the first instance; digitally delivered physician consultations arise as escalations from these contacts and were therefore treated as follow-up rather than index events. At encounter level, every digital clinic encounter in 2023-2025 was an index event, and we recorded whether a physician consultation occurred the same day and whether a further encounter was booked during it. At patient level, each patient’s first digital clinic encounter served as the index event, and follow-up was defined as the first subsequent primary care encounter occurring more than 24 hours after the index encounter that had not already been scheduled before it, within pre-specified windows of two, 14, and 30 days. Unscheduled contacts, which carry no booking record, were counted as unplanned follow-ups. Follow-up was defined without reference to the recorded diagnosis, so a subsequent encounter indicates return contact with the service rather than continued care for the index presentation. Follow-ups arranged during the index encounter itself were identified from booking date, are included in the overall rates, and are shown separately in Figure 5.

### Statistical Analysis

Demographic characteristics were compared using χ² tests for categorical variables and a two-sided Mann-Whitney U test for age, given its non-normal distribution; these comparisons treat person-years as independent observations. Comorbidity burden was compared in two ways. First, prevalence of CCI ≥ 1 was directly standardised to the age band and sex distribution of all person-years in 2023-2025. Second, logistic regression models were fitted with CCI ≥ 1 as the binary outcome and digital versus traditional user status as the exposure, adjusted for age, age², and sex: separately for each calendar year, and pooled across years with calendar year as a covariate and cluster-robust standard errors at the patient level. Results are reported as adjusted odds ratios (ORs) with 95% confidence intervals. All tests were two-tailed with a significance level of *P*<.05. Analyses were performed using Python (version 3.12.10) [16–21].

### Ethics and Approval

The study was approved by the Päijät-Häme Wellbeing Services County for secondary use of health data (decision number HA/187/07.01.04.05/2025). This study was based on secondary use of pseudonymised administrative data. In accordance with the Finnish Act on the Secondary Use of Health and Social Data (552/2019) [22] and the research permit, individual patient consent or separate ethical review was not required.

## Results

### Growth of Digital Care, 2019-2025

Service coverage declined from 39.2% in 2019 to 35.7% in 2020, and total encounters per 1,000 residents fell from 1,434 to 1,222 over the same period, consistent with the widespread suppression of non-urgent primary care encounters observed during the COVID-19 pandemic. Recovery began in 2021 coinciding with the establishment of Harjun terveys and the introduction of digital services, with coverage and utilisation subsequently exceeding pre-pandemic levels from 2021 onwards. Coverage peaked at 47.1% in 2023 and declined to 44.2% in 2025, while encounters per 1,000 continued to rise. The digital share of all primary care encounters rose from 15.2% in 2021 to 25.8% in 2025 (Table 1, Figure 1). Both the proportion of the population reached by digital services and the mean number of digital encounters per patient increased consistently across 2023-2025.

**Figure 1.**
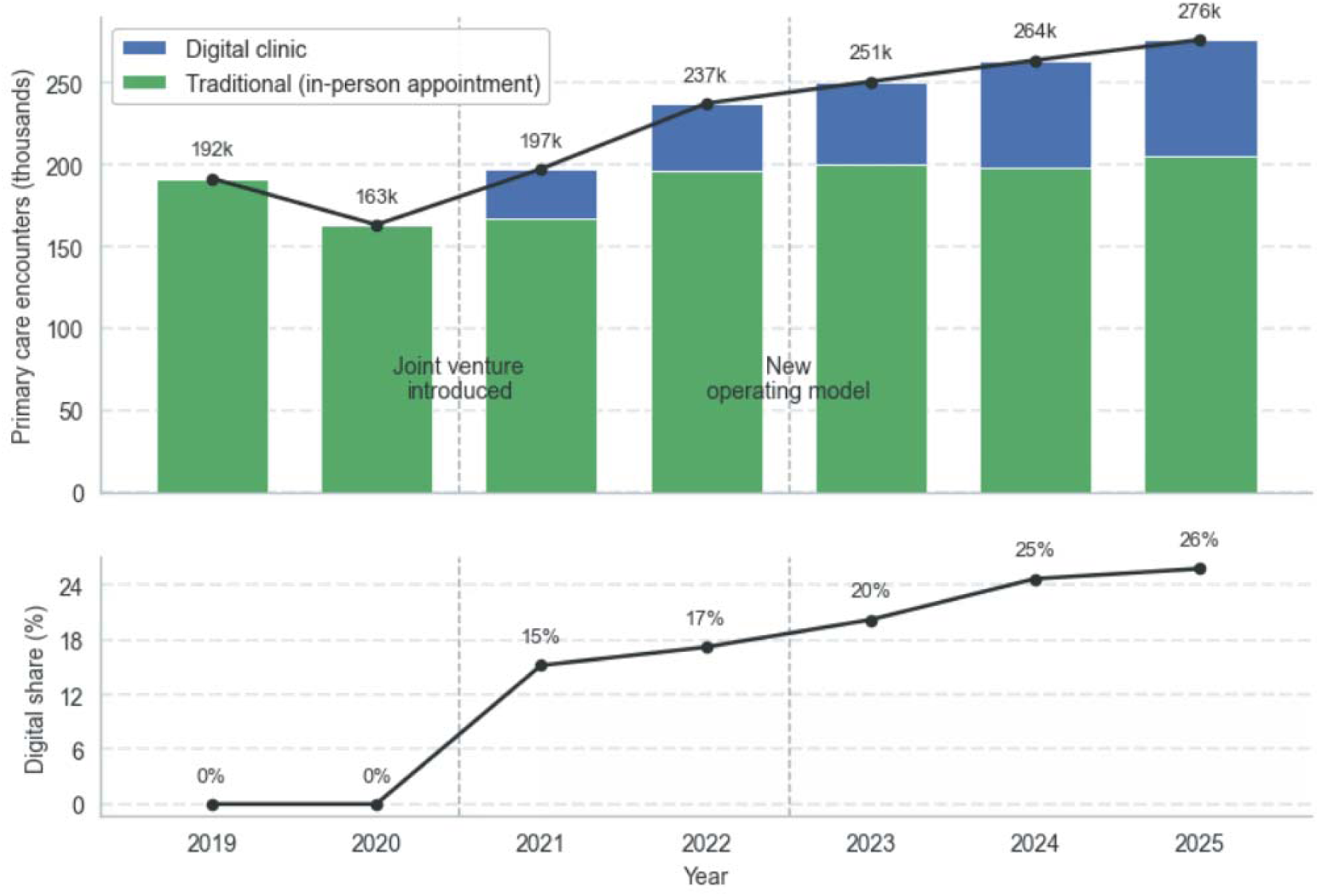
Total primary care encounters by modality, Harjun terveys, 2019-2025. The stacked bar chart shows annual encounters (in thousands) stratified by traditional encounters (in-person appointments and face-to-face physician consultations, green) and digital encounters (digital clinic encounters and digitally delivered physician consultations, blue). The line indicates total encounter volume. The lower panel displays the digital share (%) of total encounters for each year. Telephone contacts, digital documentation, and other contact types are excluded, as described in the Methods. The digital clinic was introduced in 2021, and the centralised model implemented in 2023.

**Table 1.** Annual primary care coverage (%) and encounters per 1,000 population, 2019-2025. Coverage represents the proportion of the registered population with at least one primary care encounter during the year. Encounters comprise in-person appointments at a health centre, chat-based digital clinic encounters, and physician consultations, delivered either digitally or face to face. Telephone contacts, digital documentation entries, and encounters recorded under other contact types were excluded; the rationale for each exclusion is given in Supplementary Methods 2. Dashes indicate years before digital services were introduced. Digital encounters are those recorded as digitally delivered, comprising digital clinic encounters and digitally delivered physician consultations. Population denominators were derived from national population registry data [15]; 2024 population data were used for 2025, as 2025 figures were not yet available at the time of analysis. For 2021-2022, and for evening encounters in January-April 2023, digital clinic records were held in a separate administrative system with distinct patient identifiers. Total and digital encounter counts, and therefore encounters per 1,000 and the digital share, are exact for all years and were derived by summing counts across both systems. Overall coverage for 2021-2023, and consequently the mean encounters per patient for those years, incorporate an empirical correction (φ = 0.654) for expected cross-system patient overlap derived from the fully linked 2024-2025 data; see Supplementary Methods 3. Sensitivity analyses across the full range of plausible overlap assumptions are presented in Supplementary Table S1.

| Year | Overall coverage (%) | Digital coverage (%) | Encounters per 1,000 | Digital share (%) | Mean encounters per patient | Mean digital encounters per patient |
| --- | --- | --- | --- | --- | --- | --- |
| 2019 | 39.2 | - | 1,434 | - | 3.66 | - |
| 2020 | 35.7 | - | 1,222 | - | 3.42 | - |
| 2021 | 40.1 | 9.7 | 1,479 | 15.2 | 3.69 | 0.56 |
| 2022 | 45.3 | 11.8 | 1,779 | 17.2 | 3.93 | 0.68 |
| 2023 | 47.1 | 15.3 | 1,875 | 20.2 | 3.98 | 0.80 |
| 2024 | 47.0 | 16.6 | 1,965 | 24.7 | 4.18 | 1.03 |
| 2025 | 44.2 | 18.7 | 2,058 | 25.8 | 4.66 | 1.20 |

### Demographic Profile of Digital Users

The following patient-level analyses are restricted to 2023-2025, reflecting the availability of fully linked patient-level data from the centralised digital clinic system; this restriction is described in detail in the Methods section.

Digital users were younger (mean age 33.2 vs. 55.1 years; *P*<.001), more often female (62.9% vs. 55.4%; *P*<.001), and had lower crude comorbidity prevalence (CCI ≥ 1: 13.1% vs. 33.6%; *P*<.001) than traditional users (Table 2). Much of this difference reflected age and sex composition: after direct standardisation to the age and sex distribution of all person-years in 2023-2025, prevalence was 22.8% (95% CI 22.3-23.3) among digital users and 27.5% (95% CI 27.3-27.7) among traditional users. Adjusted for age, age², sex, and calendar year, digital users had lower odds of comorbidity (OR 0.78, 95% CI 0.75-0.81; *P*<.001). Year-specific estimates were consistent.

**Table 2.**
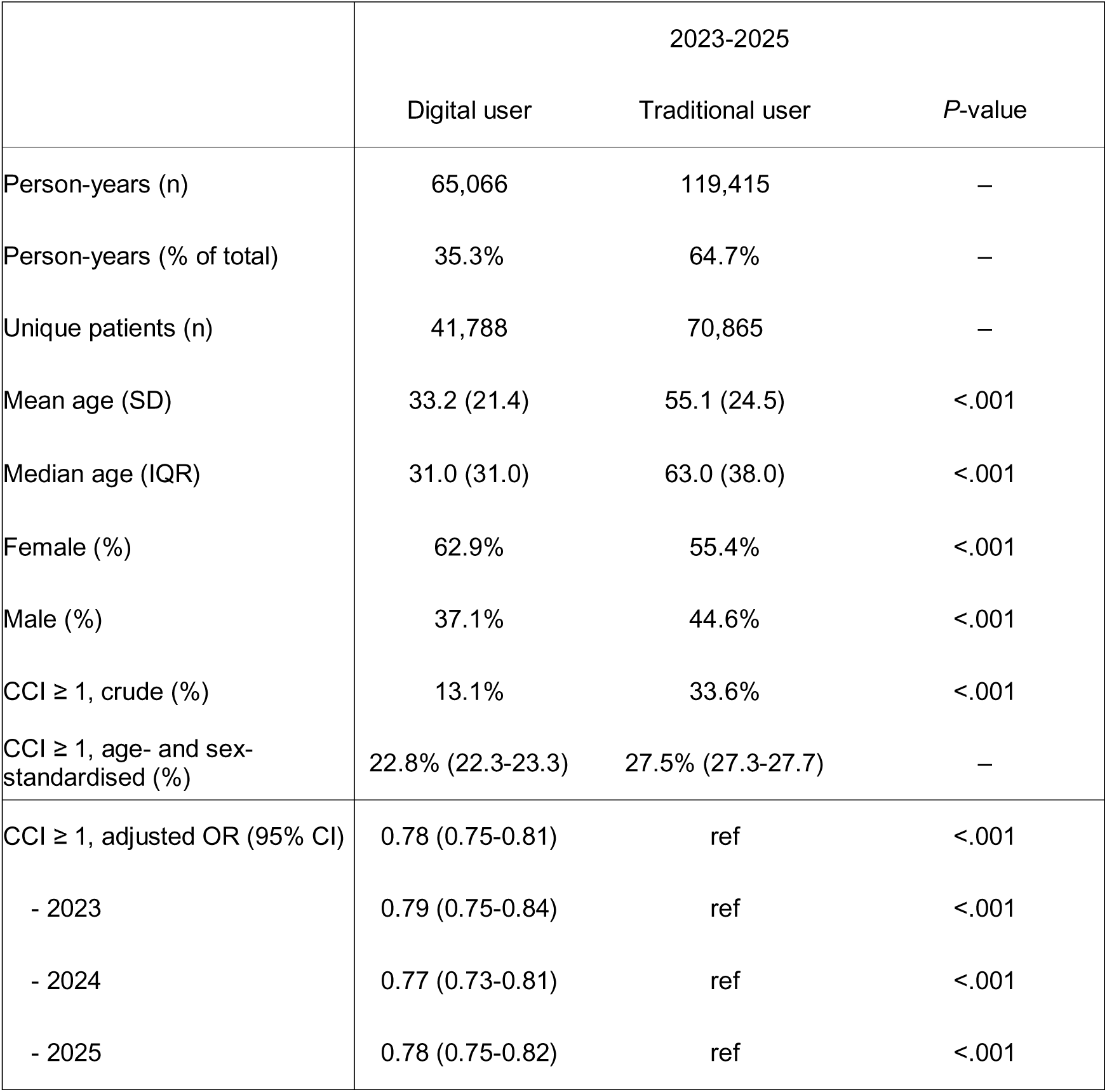
Demographic and clinical characteristics of those using healthcare services in Harjun terveys 2023-2025. Comparison of age, sex distribution, and comorbidity burden between users of digital clinic services and those using only traditional care modalities. The unit of analysis is the person-year: patients contribute one observation for each calendar year in which they had at least one primary care encounter and are classified as digital users in a given year if at least one of that year’s encounters was digitally delivered. Person-years are restricted to encounters meeting the primary care encounter definition given in the Methods; counts for 2023 are slightly lower than the corresponding coverage denominator in Table 1, which additionally incorporates patients identified only in the separate digital system. Because classification is annual, 15,676 of the 96,977 patients analysed (16.2%) appear in both columns, having used digital services in some years but not others; unique patient counts therefore sum to more than the total. Age was compared using the Mann-Whitney U test and categorical variables using the χ² test; these comparisons treat person-years as independent observations. Standardised prevalence was directly standardised to the sex distribution and age band distribution (0-17, 18-29, 30-44, 45-64, 65-74, and 75 years and over) of all person-years in 2023-2025. The pooled adjusted odds ratio is from logistic regression on CCI ≥ 1 with age, age², sex, and calendar year as covariates and cluster-robust standard errors at the patient level; year-stratified odds ratios are from models fitted separately for each year with age, age², and sex as covariates. Traditional users are the reference category in all models. There was no evidence that the association varied by year (likelihood ratio test, P=.36). CCI = Charlson Comorbidity Index.

|  | 2023-2025 |  |  |
| --- | --- | --- | --- |
|  | Digital user | Traditional user | P-value |
| Person-years (n) | 65,066 | 119,415 | — |
| Person-years (% of total) | 35.3% | 64.7% | — |
| Unique patients (n) | 41,788 | 70,865 | — |
| Mean age (SD) | 33.2 (21.4) | 55.1 (24.5) | <.001 |
| Median age (IQR) | 31.0 (31.0) | 63.0 (38.0) | <.001 |
| Female (%) | 62.9% | 55.4% | <.001 |
| Male (%) | 37.1% | 44.6% | <.001 |
| CCI $\geq 1$, crude (%) | 13.1% | 33.6% | <.001 |
| CCI $\geq 1$, age- and sex-standardised (%) | 22.8% (22.3-23.3) | 27.5% (27.3-27.7) | — |
| CCI $\geq 1$, adjusted OR (95% CI) | 0.78 (0.75-0.81) | ref | <.001 |
| - 2023 | 0.79 (0.75-0.84) | ref | <.001 |
| - 2024 | 0.77 (0.73-0.81) | ref | <.001 |
| - 2025 | 0.78 (0.75-0.82) | ref | <.001 |

Digital utilisation peaked in adolescents and young adults aged 15-29 years and declined steeply after age 30, whereas telephone and in-person utilisation increased markedly from age 65 onwards (Figure 2).

**Figure 2.**
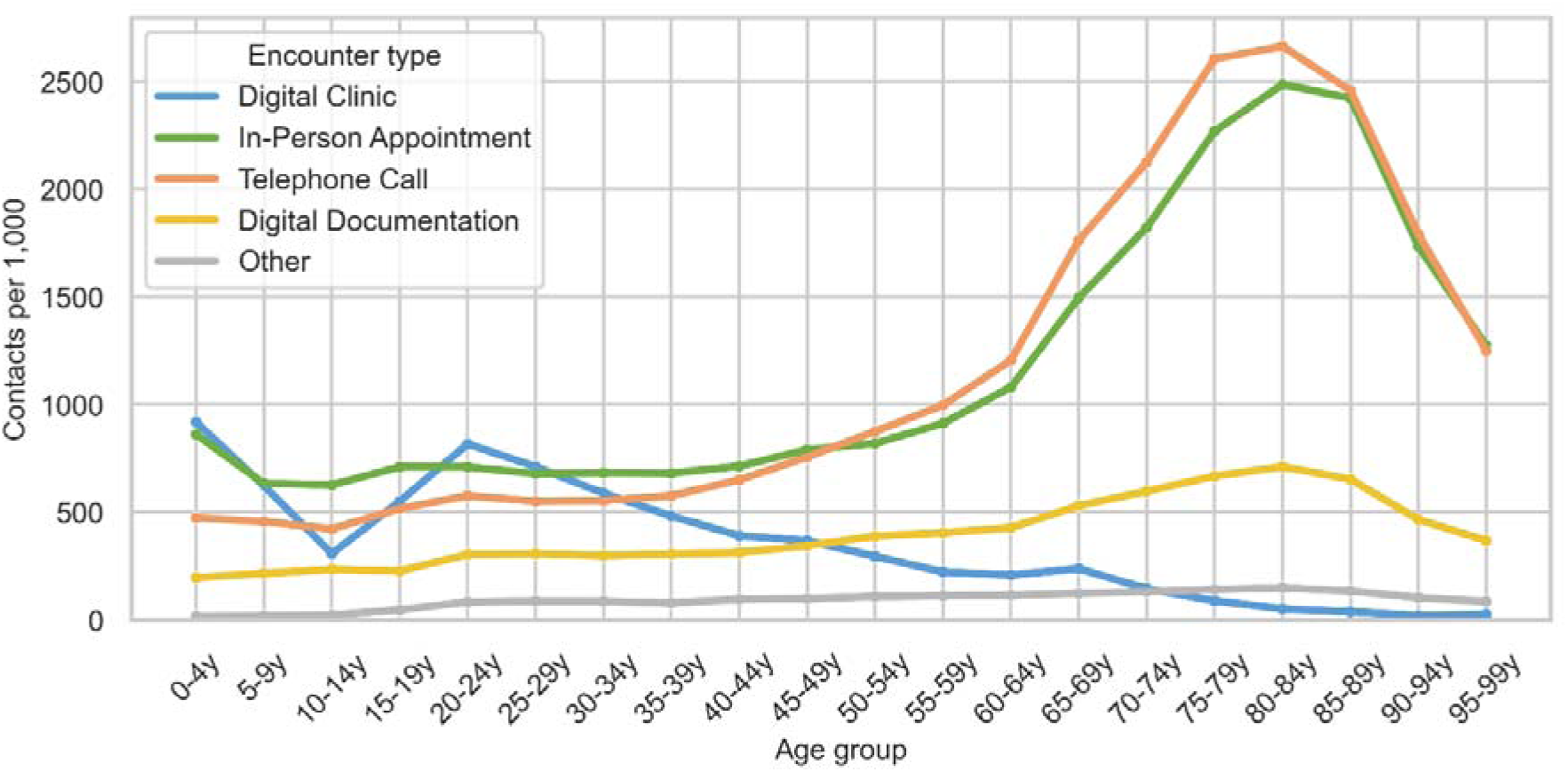
Annualised primary care encounters per 1,000 population by age group and encounter type, 2023-2025. Encounter types include digital clinic (blue), in-person appointments (green), telephone calls (orange), digital documentation (yellow), and other (grey). Unlike Table 1 and Figure 1, this figure includes telephone contacts and digital documentation, which are excluded from the primary care encounter definition; physician consultations are not shown.

### Digital Consultation Characteristics: Diagnoses and Conditions Managed

The 20 most common ICPC-2 codes collectively accounted for 46.2% of all digital encounters (n = 183,515), as shown in Figure 3. The most common reasons for encounters were respiratory infections (R74 and R83, 13.0%). Other frequent codes included general symptom consultations such as health maintenance or preventive advice (−45 and A98, 7.1%), various skin symptoms (S29 and S21, 5.2%), and throat or eye-related symptoms (R21, F03, and F29, 6.4%). Notably, administrative interactions related to prescription renewal were also featured (−50, 1.9%).

**Figure 3.**
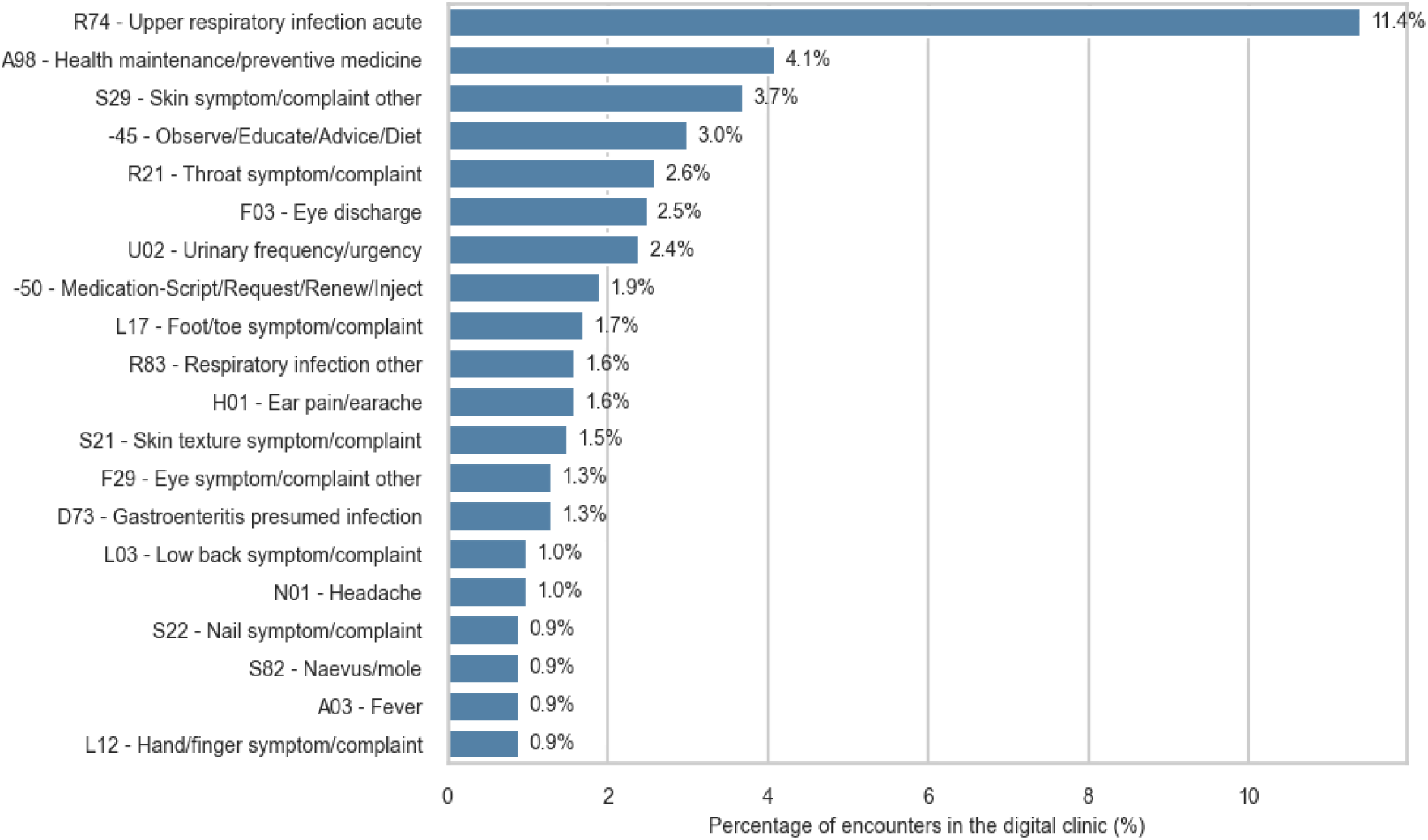
Most common ICPC-2 diagnosis categories in the digital clinic (2023-2025). The 20 most frequent ICPC-2 codes recorded in digital encounters, expressed as a percentage of all 183,515 digital encounters (digital clinic encounters and digitally delivered physician consultations) recorded between 2023 and 2025.

Figure 4 shows the 10 most frequent ICD-10 diagnoses recorded in physician-level digital consultations, expressed as a percentage of all digital encounters (n = 183,515). The most common were conjunctival infections (H10.x combined, 1.6%), acute cystitis (N30.0, 1.2%), and prescription renewals (Z76.0, 1.2%). Additional diagnoses reflected a typical profile of low-complexity primary care issues, including cellulitis, dermatitis, and unspecified acute upper respiratory infection.

**Figure 4.**
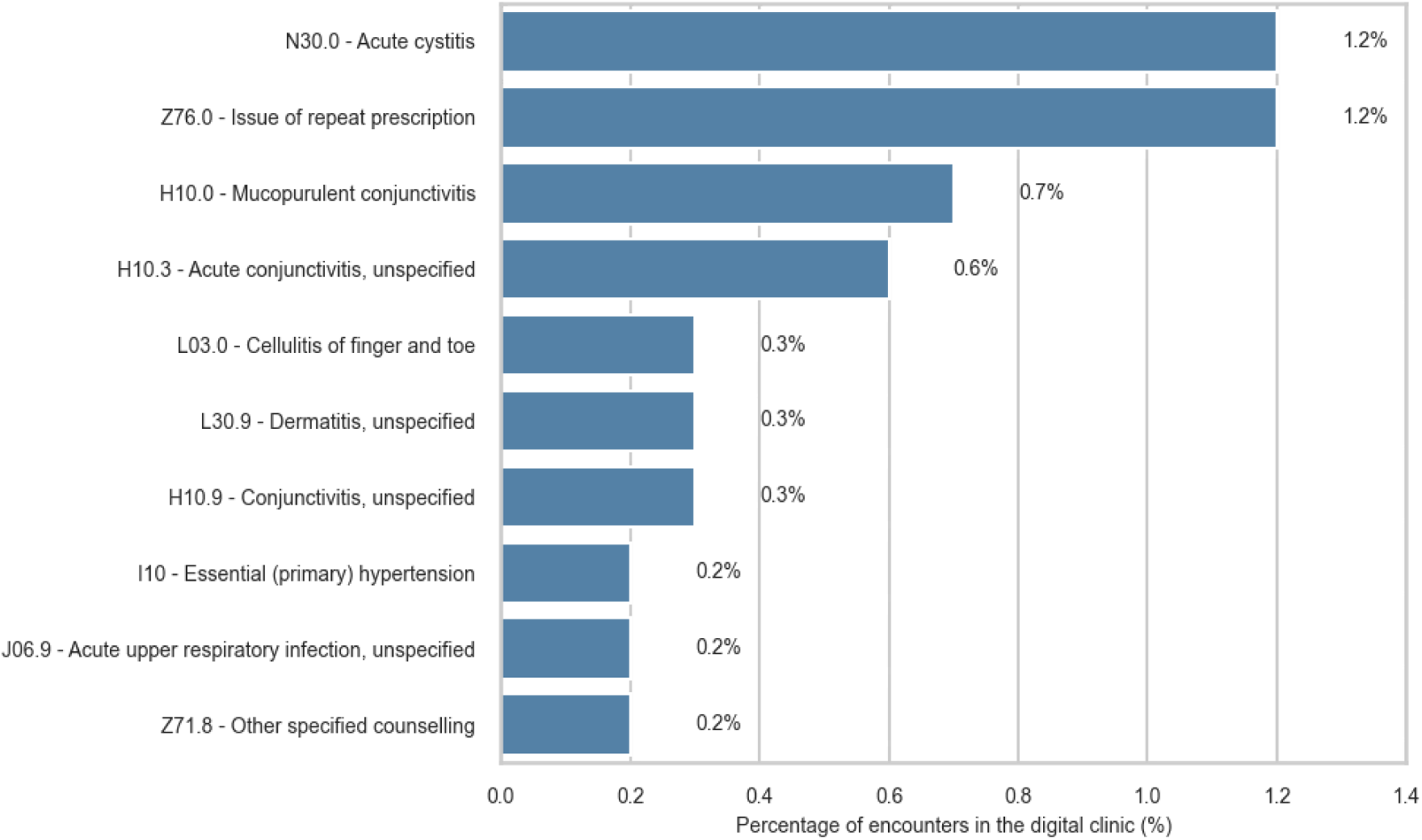
Most common ICD-10 diagnosis categories in the digital clinic (2023-2025). Top 10 ICD-10 codes recorded during digital consultations with a physician, following escalation from the nurse encounter at the digital clinic. Percentages represent the share of all 183,515 digital encounters (digital clinic encounters and digitally delivered physician consultations) recorded between 2023 and 2025, not the share of physician consultations.

### Follow-up Analysis

To evaluate care continuity following nurse-led digital encounters, we analysed subsequent primary care encounters over a 30-day window for the period 2023-2025. The encounter-level analysis comprised all 154,695 digital clinic encounters recorded between 2023 and 2025. On the same day as the digital encounter, 18.0% (n = 27,820) involved direct escalation to a physician consultation, and 14.0% (n = 21,591) resulted in a new subsequent encounter being booked for a later date.

Time to first subsequent contact was then examined at patient level, taking each patient’s first digital clinic encounter in 2023-2025 as the index event (n = 40,724 patients) and counting any subsequent primary care encounter occurring more than 24 hours later that had not already been scheduled before the index encounter. Contacts were counted irrespective of recorded diagnosis and therefore represent all-cause return contact. Across all modalities, 13.1% of patients had a subsequent contact within two days, 32.6% within 14 days and 42.4% within 30 days; the remaining 57.6% had no recorded contact within 30 days. Figure 5 presents the cumulative incidence by encounter type. A repeat digital clinic encounter was the most common form of return contact (6,331 of 17,283 first contacts), followed by in-person appointments (3,883), telephone contacts (3,402), digital documentation entries (2,253) and physician consultations (1,130).

**Figure 5.**
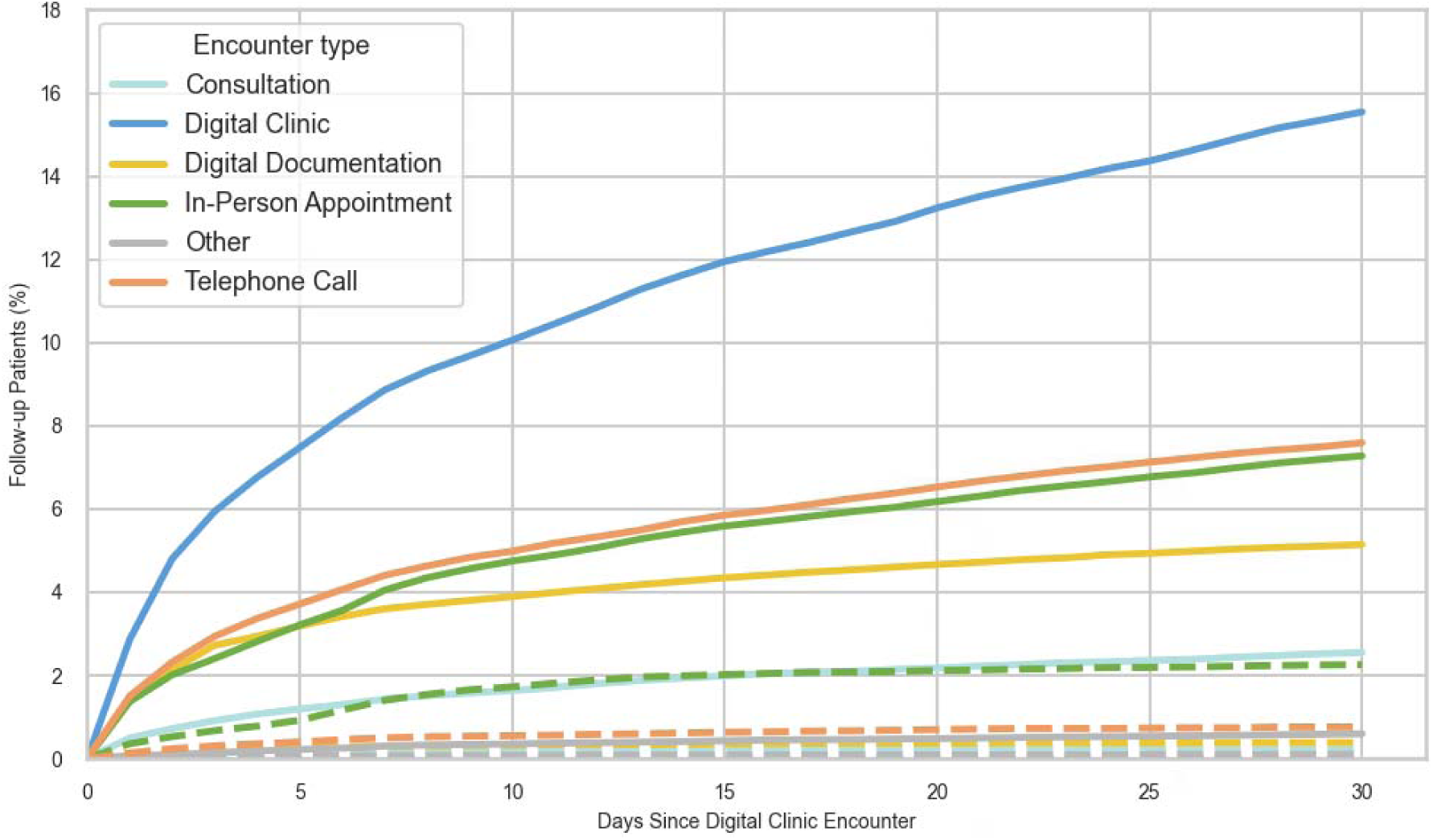
Cumulative follow-up by encounter type over 30 days following an encounter with the digital clinic. Solid lines show contacts that were unscheduled or booked after the index encounter; dashed lines show, separately, those arranged during the index encounter itself. Encounters already scheduled before the index encounter are excluded from both. Repeat digital clinic encounters carry no booking record and therefore appear only as solid lines. Contacts are counted irrespective of recorded diagnosis.

Most return contacts were not planned at the index encounter: 1,519 of the 17,283 first contacts within 30 days (8.8%) had been booked during it, ranging from 23.7% of in-person appointments to 9.1% of telephone contacts and 8.1% of physician consultations. The most common presenting reasons for these pre-booked follow-ups were skin lesions (naevus/mole), lacerations, acute upper respiratory tract infections, diabetes, and sleep disturbances. Repeat digital clinic encounters are unscheduled by design and carry no booking record, so they appear entirely as unplanned contacts.

Figure 6 presents a Sankey diagram of care pathways over the first 7 days after a digital clinic encounter, covering all planned and unplanned utilisation. Approximately 60% of patient trajectories ended within the digital channel, either directly following the nurse encounter or after a physician consultation. A physician consultation was an intermediate step in 23% of all encounters, after which patients either concluded the episode or went on to in-person, new digital, telephone, or other follow-up encounters.

**Figure 6.**
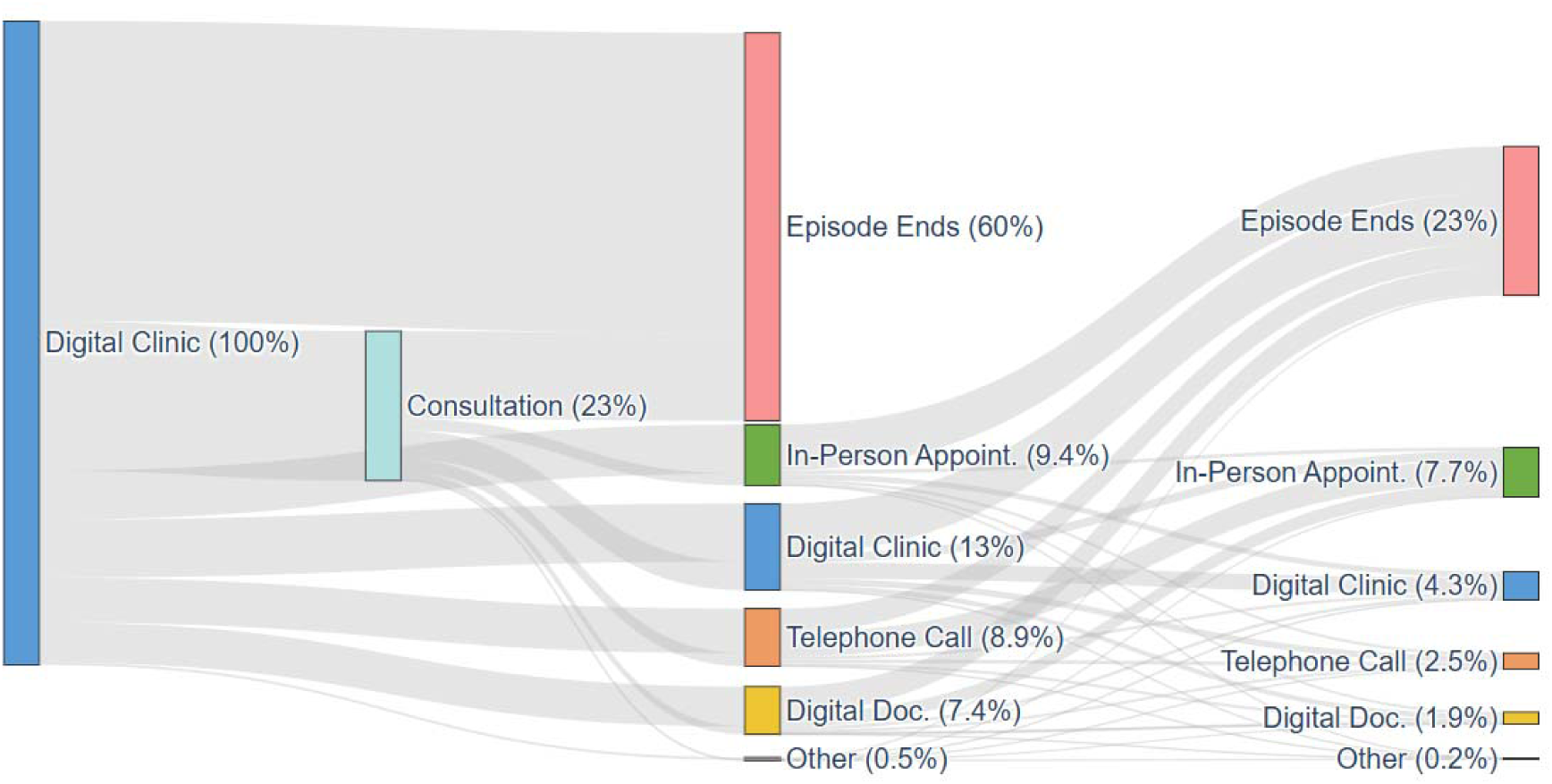
Sankey diagram of healthcare utilisation after a digital clinic encounter, 2023-2025. Flow widths correspond to the proportion of patients at each step. The diagram covers the first 7 days after the encounter, and sequences are truncated at three steps to aid interpretation.

## Discussion

This study characterised the utilisation, patient profile, clinical scope, and care pathways of a 24/7 chat-based digital clinic integrated into publicly funded primary care in Finland. Digital encounters grew from 15.2% of all primary care encounters at launch to approximately 26% by 2025, and were predominantly used for acute, low-complexity presentations, most of which were concluded without a planned follow-up appointment. Because the digital clinic was introduced concurrently with broader joint-venture organisational reforms, and because the 2020 contraction in care-seeking was followed by post-pandemic recovery, its independent contribution to system-level changes in coverage cannot be established with the present design.

Patterns of digital clinic adoption were consistent with those reported in prior European studies, with higher uptake among younger adults and women [9–11]. Older adults and patients with a higher burden of chronic disease continued to rely primarily on traditional, non-digital care. The digital channel appears to attract patients with acute, episodic care needs rather than those with substantial chronic disease burden, consistent with the predominance of minor acute diagnostic codes in the clinical content analysis.

A substantial part of the crude difference in comorbidity between digital and traditional users was attributable to age and sex: standardising to a common age and sex distribution reduced a 20.5 percentage point crude gap to 4.7 points. A clear difference nonetheless persisted after adjustment (OR 0.78, 95% CI 0.75-0.81) and was stable across the three years. Digital users of equivalent age and sex therefore carry a lower comorbidity burden than those using traditional services alone, by roughly a fifth on the odds scale. This likely reflects both clinical appropriateness, with patients managing complex chronic conditions continuing to rely on in-person care, and selection effects, whereby healthier individuals within each age stratum adopt digital services more readily.

Approximately 86% of digital encounters did not result in a follow-up appointment being booked at the time of the digital encounter, consistent with a case mix of acute, low-complexity presentations that can often be concluded in a single contact. Where patients did contact the service again, they largely stayed in the digital channel: a repeat digital clinic encounter was the most common first return contact (6,331 of 17,283), while an in-person appointment was the first return contact for 9.5% of index patients within 30 days. Because encounters cannot be linked to a clinical problem in these data, these are all-cause contact rates: they describe how patients continued to use the service, not whether the index presentation was resolved. On that basis they are broadly in line with Glock et al. [11], who reported that 43.6% of eVisits to a Swedish all-virtual primary care unit were followed by any healthcare encounter within 14 days, and with Lakoma et al. [9], who found that approximately 70% of problems were resolved at the digital clinic. Both studies identified acute cystitis and urinary tract infections among the most common drivers of in-person follow-up, presentations now frequently managed within the digital pathway. In the present study, digital encounters were frequently followed by physician consultations where prescription authority was required, reflecting the intended nurse-led design in which nurses are the initial point of contact and escalate when clinically indicated.

Among the minority of encounters that progressed to an in-person visit, conditions requiring physical examination were overrepresented, including skin lesions such as naevi, lacerations, and respiratory conditions in which pneumonia was suspected. This reflects case selection rather than duplication of care: presentations suited to remote assessment were largely resolved within the digital channel, while those requiring examination were identified and redirected, the digital encounter serving much the same triage function as a telephone encounter.

Lower engagement among older adults may reflect the threshold to begin using the service (device access, application installation, and electronic identification), usability barriers once in use, or preferences for in-person interaction. Most digital encounters were episodic, and few patients with high comorbidity burden used the digital pathway for routine chronic care follow-up. Expanding digital services to include selected chronic care tasks, such as remote monitoring, is a promising direction for future development [23]. Evidence indicates that digital services can improve access for specific populations [24,25]. The lower uptake among older adults in this setting points to the core implementation challenge: ensuring that digital expansion does not widen access gaps. In line with previous recommendations [26–29], hybrid models that preserve in-person options, build users’ digital skills, and offer personalised support (including remote assistance), together with patient-centred platform design with intuitive interfaces and clear clinical guidance, can help broaden inclusion [29,30].

### Limitations

This study relied on routinely collected health service data, which carries inherent limitations. Some outcomes like clinical resolution or patient satisfaction were not directly measured; we inferred resolution from return-contact rates, which are limited in two directions. Encounters could not be linked to a clinical episode or problem, so a subsequent contact cannot be attributed to the index presentation and these rates describe all-cause return contact, overstating the proportion of episodes that went unresolved. Conversely, care sought outside the study system, including occupational health and private providers, is not observed, so resolution within the digital channel may be overstated. There is also potential misclassification in diagnostic coding. Telephone contacts were excluded from annual totals because of discontinuities in how they were recorded (Supplementary Methods 2), although they are retained as a possible follow-up modality in the pathway analyses. Since this was an observational study within one wellbeing services county, generalisability may be limited; factors unique to Päijät-Häme (such as the specific app used or population characteristics) might affect results. Additionally, our comparisons between digital and traditional users are subject to confounding, as the groups differ by age and health status by design.

The φ-correction applied to coverage estimates for 2021-2023 rests on assumptions that are not fully verifiable and most plausibly overcorrects: if early-phase digital users relied on the digital channel more exclusively than users in the mature system, the true φ was lower in those years and coverage for 2021-2022 is underestimated. Across the full range of plausible φ values (0 to 1), coverage for 2021 ranges from 36.8% to 46.5% and for 2022 from 41.2% to 53.0% (Supplementary Table S1), so the direction of the trend is robust even though the precise estimates carry uncertainty.

Comorbidity was taken from a single population registry extract rather than measured as of each encounter, so the same Charlson Comorbidity Index value was applied to all person-years contributed by a given patient.

Attribution of effects should be cautious. Concurrent system improvements under the joint venture mean that attributing all observed changes to the digital channel risks overstatement. As an observational study, we cannot establish causality.

### Conclusions

Digital encounters through the Päijät-Sote application grew to account for approximately 26% of all primary care encounters by 2025, predominantly serving younger adults with acute, low-complexity presentations, most of which were concluded without a planned follow-up. Escalation to physician consultation or in-person care followed a pattern consistent with the nurse-led triage model.

Digital service use was concentrated among younger patients. After accounting for age and sex, digital users had lower odds of comorbidity than traditional users (OR 0.78, 95% CI 0.75-0.81), a smaller difference than the crude prevalence gap suggests but a consistent one across all three years. Older adults continued to rely primarily on traditional care. As digital services expand within publicly funded systems, equity-conscious implementation, including hybrid models, digital literacy support, and accessible platform design, will be essential to ensure that efficiency gains do not come at the cost of inclusive access.

## Declaration Statements

### Data Availability

All data generated or analysed during this study are presented within the published article and its supplementary materials. Patient-level data are not publicly available due to privacy and data protection regulations.

### Code Availability

The underlying code for this study is available at https://github.com/achdahlberg/coverage-utilisation-digital-clinic-2026.

## Acknowledgements

The authors gratefully acknowledge the support provided by Mehiläinen Oy, Harjun terveys oy, and the Päijät-Häme Wellbeing Services County. AD acknowledges Päijät-Hämeen Lääkäriseura. ChatGPT [31] was used to assist in improving the clarity and coherence of the manuscript. All content was thoroughly reviewed, edited, and approved by the authors, who take full responsibility for the accuracy and integrity of the final work.

## Authors’ Contributions

**A.D.:** Conceptualisation, Data curation, Formal analysis, Investigation, Methodology, Project administration, Software, Validation, Visualisation, Writing - original draft, Writing - review & editing. **T.K.:** Conceptualisation, Resources, Supervision, Validation, Writing - review & editing. **S.J.:** Conceptualisation, Methodology, Resources, Software, Supervision, Validation, Writing - review & editing. **P.O.:** Conceptualisation, Funding acquisition, Project administration, Resources, Supervision, Writing - review & editing.

