## Supplementary Information for "Coverage, Utilisation and Users of a Chat-Based Digital Primary Care Clinic in Publicly Funded Healthcare: A Registry-Based Observational Study in Finland"

### Supplementary Methods

#### 1. Setting and organisational context

Harjun terveys was established in January 2021 as a partnership between the regional joint municipal authority for health and social services, whose responsibilities transferred to the Päijät-Häme Wellbeing Services County in 2023, and a private healthcare company. The venture was created to address sustained shortfalls in local service capacity relative to patient demand and delivers outpatient medical and dental services; dental encounters were excluded from the present analysis. In some Finnish municipalities, parts of public service delivery are outsourced to private healthcare providers. The catchment population is urban-rural mixed, with an age distribution slightly older than the Finnish national average [1]. The digi-physical service model, in which a 24/7 chat-based digital clinic operates alongside telephone and in-person appointments, is intended to reduce appointment delays and improve access to same-day or rapid assessment.

The observation period spans three phases of organisational and digital reform: two pre-implementation years (2019-2020); the establishment of the joint venture and the introduction of system-wide digital services (2021-2022); and the operation of a centralised digital clinic under a standardised model (2023-2025).

#### 2. Encounter definition, exclusions and record flow

The initial extract comprised 2,902,707 primary care encounters recorded between 1 January 2019 and 31 December 2025. After deduplication, exclusion of encounters with unknown or out-of-scope profession codes (including dental encounters), and removal of records with missing date of birth, 2,796,395 encounters remained, a net reduction of 106,312 records (3.7% of the original entries).

Of these 2,796,395 records, 1,505,993 met the primary care encounter definition given in the Methods. A further 74,845 encounters recorded in the separate digital system bring the total to 1,580,838 encounters, which form the basis of the coverage and utilisation analyses reported in Table 1 and Figure 1.

Four categories present in the extract were excluded. Health promotion and preventive services, such as vaccinations and maternity or child health clinic visits, do not represent demand-driven care seeking. Digital documentation entries record clinical documentation rather than a patient contact. A residual category of encounters could not be classified to a defined modality. Telephone contacts were excluded because the way they were recorded changed repeatedly over the observation period, in ways unrelated to the volume of care delivered: recorded telephone volume rose by 62% between 2020 and 2021, coinciding with the establishment of the joint venture, and by a further 44% between 2024 and 2025, with intervening years showing no comparable movement. Telephone contacts are nonetheless retained in the follow-up analyses, where they are examined as one of several possible follow-up modalities rather than counted towards annual totals.

#### 3. Cross-system linkage and the overlap correction

For 2021-2022 and evening encounters in January-April 2023, digital clinic records were held in a separate administrative system with distinct patient identifiers that could not be linked to the primary EHR. This introduced a methodological distinction between encounter-level and patient-level metrics. Total and digital encounter counts, and therefore encounters per 1,000 residents and the digital share, were computed by summing counts across both systems and are exact for all years. Patient-level coverage estimates required correction, as direct addition of unique patient counts across systems would risk double-counting individuals who used both modalities within the same year. Mean encounters per patient depends on the unique patient count and is therefore subject to the same correction as coverage.

An empirical overlap fraction φ was derived from the fully linked 2024-2025 data, in which digital and traditional encounter records share a common patient identifier. For each linked year, φ was defined as the proportion of unique digital users who also had at least one traditional encounter during that calendar year; the mean across both years (φ = 0.654) was applied to the unlinked period. Corrected coverage was estimated as

$$N_{covered} = N_{EHR} + N_{digital} \times (1 - \varphi)$$

where $N_{EHR}$ is the number of unique patients in the primary EHR, $N_{digital}$ is the number of unique patients in the separate digital system, and $N_{digital} \times (1 - \varphi)$ represents the expected number of digital patients not already captured in the EHR.

Digital coverage for 2021 and 2022 was derived directly from the unique patient count in the separate digital system, as no digital encounters were recorded in the primary EHR in those years. For 2023, digital coverage combines unique patient counts from both systems; because the two could not be linked, patients who used both the centralised digital clinic and the separate evening service during that year are counted twice, and the 2023 figure is therefore a slight overestimate. The separate digital system contributed 3,823 evening encounters from 2,877 unique patients in January-April 2023, a small fraction of total annual volume.

This approach assumes that the propensity for overlap between digital and traditional service use was similar in 2021-2023 to that observed in 2024-2025. Early adopters of digital services in newly launched systems may be more likely to use the digital channel as their primary or exclusive point of contact, implying a lower true φ in those years and hence an over-large adjustment that would underestimate coverage for 2021-2022. Sensitivity analyses across the full range of plausible φ values (0 to 1) are presented in Supplementary Table S1.

### Supplementary Tables

***Supplementary Table S1 Sensitivity of annual service coverage estimates for 2021-2023 to alternative values of the empirical overlap fraction φ***. *φ = empirical overlap fraction, defined as the proportion of unique digital clinic users who also had at least one traditional primary care encounter in the same calendar year. φ = 0.0: upper bound, assumes zero overlap between systems (all digital patients are genuinely new unique individuals not captured in the EHR). φ = 1.0: lower bound, assumes complete overlap (all digital patients already appear in the EHR). Primary estimate uses φ = 0.654, derived as the mean overlap fraction across 2024 and 2025, the years in which both digital and traditional encounter records share a common patient identifier within the EHR.* $Corrected\ n = unique\ patients\ in\ the\ primary\ EHR + unique\ patients\ in\ the\ separate\ digital\ system \times (1 - \varphi);$ $corrected\ coverage\ (\%) = corrected\ n/population\ \times 100.$

*Shaded rows indicate the primary analysis estimate. Population denominators derived from national population registry data (Official Statistics of Finland, Population structure; https://stat.fi/tilasto/vaerak); 2024 data used as proxy for 2025.*

| Year | φ value | Corrected n | Corrected coverage (%) | Notes |
|---|---|---|---|---|
| 2021 | 0.0 | 62,069 | 46.5% | Upper bound (zero overlap assumed) |
| 2021 | 0.2 | 59,470 | 44.6% | |
| 2021 | 0.4 | 56,872 | 42.6% | |
| 2021 | 0.6 | 54,273 | 40.7% | |
| **2021** | **0.654** | **53,572** | **40.1%** | **Primary estimate** |
| 2021 | 0.8 | 51,675 | 38.7% | |
| 2021 | 1.0 | 49,076 | 36.8% | Lower bound (full overlap assumed) |
| 2022 | 0.0 | 70,759 | 53.0% | Upper bound (zero overlap assumed) |
| 2022 | 0.2 | 67,604 | 50.7% | |
| 2022 | 0.4 | 64,449 | 48.3% | |
| 2022 | 0.6 | 61,294 | 45.9% | |
| **2022** | **0.654** | **60,442** | **45.3%** | **Primary estimate** |
| 2022 | 0.8 | 58,139 | 43.6% | |
| 2022 | 1.0 | 54,984 | 41.2% | Lower bound (full overlap assumed) |
| 2023 | 0.0 | 64,839 | 48.5% | Upper bound (zero overlap assumed) |
| 2023 | 0.2 | 64,264 | 48.0% | |
| 2023 | 0.4 | 63,688 | 47.6% | |
| 2023 | 0.6 | 63,113 | 47.2% | |
| **2023** | **0.654** | **62,957** | **47.1%** | **Primary estimate** |
| 2023 | 0.8 | 62,537 | 46.7% | |
| 2023 | 1.0 | 61,962 | 46.3% | Lower bound (full overlap assumed) |


[1] Official Statistics of Finland. Population structure 2025. Available from: https://stat.fi/tilasto/vaerak
